# Life course shaping of brain ageing: the SHARE blood biomarker study

**DOI:** 10.64898/2026.05.17.26353413

**Authors:** Gindo Tampubolon, Guannan Li

## Abstract

**Background:** Evidence from many countries shows that later life cognitive health is shaped by childhood poverty. However, whether it is associated with neurodegenerative biomarkers measured in population settings remains unclear.

**Methods:** We conducted a pooled analysis of 5,473 adults aged ≥50 years from Denmark, Sweden and Germany participating in Wave 6 (2015) of the Survey of Health, Ageing and Retirement in Europe. Neurodegenerative biomarkers (neurofilament light chain, glial fibrillary acidic protein and phosphorylated tau) were assayed from dried blood spots. Childhood poverty was constructed as a latent variable from retrospective life histories. Weighted Poisson regression models estimated associations adjusting for age, sex, education, marital status and wealth in later life. Marginal predictions along age and across country were derived.

**Results:** Childhood poverty was strongly associated with higher NfL concentrations (*β* = 1.66, *p* < 0.001), but not with GFAP or p-tau217. Predicted values indicated substantially elevated NfL among the childhood poor (10.3 pg/mL vs 2.0 pg/mL for the non-poor). Age profiles showed widening disparities: the childhood poor in midlife exhibited higher NfL levels than the oldest old who grew up not poor. No consistent differences were observed for GFAP or p-tau217. Findings were robust and similar across all three countries with different histories and health systems.

**Conclusions:** Childhood poverty is associated with markedly elevated levels of NfL in later life, suggesting long-term neuroaxonal injury consistent with life course shaping of brain health. Moreover, the evidence implies substantial acceleration of neurobiological ageing. These findings emphasise the importance of early-life interventions for brain health in ageing populations.

**Key points:**

- Childhood poverty is associated with higher concentrations of neurofilament light chain (NfL) in later life.
- No corresponding association is observed for glial fibrillary acidic protein or phosphorylated tau at threonine 217.
- Differences in NfL widen with age, indicating cumulative life-course effects on neuroaxonal integrity.
- The childhood poor in midlife record NfL concentrations exceeding those of the oldest old who grew up not poor.
- Dried blood spot sampling is a feasible method for measuring neurodegenerative biomarkers in population-based surveys.

## Introduction

Cognitive impairment and dementia are among the most consequential threats to the capacity to live independently and participate freely in society, driving disability, long-term care demand, and social inequalities.^1^ Emerging evidence now shows that later life cognitive function is not determined by later life circumstances only. It is shaped through the life course, beginning in childhood.^2^ Cross-national analyses using ongoing longitudinal ageing studies on both sides of the Atlantic and Asia have demonstrated that childhood poverty continues to mark later life function across diverse outcomes and settings.^3–5^ Childhood poverty is associated with higher probability of sarcopenia, depression and frailty as well as lower cognitive function across 28 countries, even after accounting for youth illness, education, wealth and other adult factors.^2,6^ Another multinational study across 32 wealthy and developing nations shows that childhood poverty is associated with lower healthy ageing, a multidimensional construct spanning cognition, mobility, sensory function, vitality, activities of daily living and mental health.^7^ These findings imply that responding to population ageing requires earlier interventions than those in later life alone: inequalities in later life functionings have roots in childhood material conditions.

Recent work has also progressed from documenting associations to identifying plausible mechanisms. Biological embedding offers one pathway through which childhood material conditions “get under the skin” and shape later life outcomes.^8^ Epigenetic ageing (using methylation-based measures) has been advanced as a mechanism linking early life poverty to later life health and functioning, and this mechanism has found some support in the US Health and Retirement Survey (HRS) venous blood study.^2^ This mechanistic direction strengthens the case for integrating social epidemiology with biological measurement, enabling life course research to move beyond distal cognitive outcomes toward brain ageing. A key step is to examine whether childhood poverty is associated with neurodegeneration, measured through blood-based biomarkers that reflect axonal injury, astrocytic activation, and Alzheimer’s-related tau pathology.^3^ However, population-based biomarker studies face practical constraints: venous blood collection is costly and logistically demanding, especially at scale and outside clinical settings.^5^ Dried blood spots (DBS) offer a scalable alternative: they are minimally invasive, cheaper to collect, and feasible under routine survey conditions.

The Survey of Health, Ageing and Retirement in Europe (SHARE) biomarker study provides a unique take on this step.^9^ SHARE is a large population-based longitudinal study explicitly designed to investigate health, economic and social circumstances over the life course in the context of population ageing.^2,6,7,10^ In SHARE Wave 6 (2015), DBS were collected in multiple countries under survey conditions, enabling measurement of biomarkers relevant to brain ageing. In the SHARE biomarker study, phosphorylated tau at threonine 217 (p-tau217), glial fibrillary acidic protein (GFAP) and neurofilament light chain (NfL) were analysed in a subsample from Denmark, Germany and Sweden, demonstrating the feasibility of measuring these neurodegenerative biomarkers from DBS material collected outside clinical settings.

Using neurodegenerative biomarkers collected with either method, studies on both sides of the Atlantic and South Asia show that higher levels of both GFAP and NfL are consistently related with cognitive impairment and dementias in older adults. Other neurodegenerative biomarkers such as phosphorylated tau and amyloid-*β* ratio are less so.^3–5^ Building on the external validity of these biomarkers, we then use the life course shaping of health thesis (which has been tested on frailty and healthy ageing) and the biological embedding literature to investigate the association between childhood poverty and later life neurodegeneration as measured by concentrations of NfL, GFAP, and p-tau217 in three SHARE countries (Denmark, Sweden and Germany).

This study has three aims: (1) to estimate the life course associations between childhood poverty (obtained with retrospective life histories) and concentrations of neurodegenerative biomarkers in later life, specifically NfL, GFAP and p-tau217; (2) to ascertain the feasibility of obtaining such estimates from materials obtained by the cheaper method of dried blood spot which can be deployed in population surveys outside clinical settings; and (3) to assess one of the ways that childhood material conditions get under the skin and affect cognitive function.

## Methods

### Study design and setting

We conducted a pooled, cross-sectional analysis of the SHARE biomarker study, which collected dried blood spot (DBS) samples during Wave 6 in 2015. Börsch-Supan and colleagues (2026) describe the DBS collection, laboratory assays, and post-collection adjustments implemented to support cross-country comparability in a non-clinical survey environment.^9^ DBS eligibility in SHARE Wave 6 was restricted to longitudinal households (respondents who had previously participated in any SHARE wave and their partners) and required written informed consent. For neurodegenerative biomarkers, assays were performed in 2025 on DBS material available from respondents in Denmark, Sweden, and Germany. Our pooled analytic sample comprised *N* = 5, 473 respondents with DBS-based neurodegenerative biomarker data.

### DBS collection, transport, storage, and weights

During the in-home Wave 6 interview, non-fasting capillary finger-prick blood was spotted onto DBS cards by trained interviewers and mailed to the SHARE Biobank (University of Southern Denmark, Odense) for freezer storage at −23°C until analysis and thereafter. Because DBS participation and analyzability can be selective, SHARE constructed DBS-specific weights which we used for regression analyses to reduce bias due to eligibility/consent/analyzability.

### Outcomes: neurodegenerative biomarkers

Outcomes were concentrations of neurofilament light chain (NfL), glial fibrillary acidic protein (GFAP), and phosphorylated tau at threonine 217 (P-tau217). NfL reflects neuro-axonal injury and neurodegeneration; GFAP reflects astrocyte activation and neuroinflammation; P-tau217 is associated with Alzheimer-type pathology. Laboratory extraction, assay procedures and detection limits are presented elsewhere.^9^

### Exposure: childhood poverty from retrospective life histories

Childhood conditions in SHARE are collected in retrospective life history interviews. Retrospective accounts of childhood material conditions are widely recognised to be error-laced, raising the risk of unsafe inference. Per prior life-course research, we operationalised childhood poverty as a latent construct indicated by multiple retrospective items rather than relying on a single recalled measure.^11,12^ This latent construct approach has been used in cross-country life course studies to reduce measurement error and bias and to allow comparison across settings where indicators differ, but the underlying construct of childhood poverty is common.^2,6,7,13^

### Covariates

All models adjusted for the following covariates: age (years), sex, education (college vs less than college), marital status (single; married/partnered; separated/divorced/widowed), wealth quartile (bottom quartile vs higher), and country (Denmark, Sweden, Germany).

### Statistical analysis

Because biomarker concentrations are naturally positive, and have very long right tail, we fitted Poisson models to each biomarker (NfL, GFAP, P-tau217) as the dependent variable, using weights supplied by SHARE. Country samples were pooled. To ease interpretation, we then draw marginal or predicted concentrations along age, distinguishing the childhood poor from non-poor, for each outcome and each country. Because childhood poverty is a derived, not observed, latent construct, standard error adjustments are applied.^14,15^ Model estimation is done in Latent Gold Syntax 6.1 and significance is set at 5%.

### Ethics and data access

SHARE DBS collection was approved by ethics committees in all participating countries and by the Ethics Council of the Max Planck Society; written informed consent was obtained and could be revoked. The University of Manchester exempted the investigation from full ethical review as it uses publicly available deidentified secondary datasets. The DBS biomarker module is available via SHARE to registered researchers (www.share-project.org).

## Results

The pooled analytic sample (Table 1) comprised 5,473 older adults from Germany (37%), Sweden (29%), and Denmark (33%). Mean age was 66 (SD 9) years, with 54% women. Nearly 8% of participants were classified as having experienced childhood poverty based on retrospective life history data. Compared to those not poor in childhood, the childhood-poor subgroup had similar age (mean 66) and sex distribution (54% female) but slightly lower educational attainment (36% vs 37% college-educated) and comparable marital status. Baseline biomarker levels were in the expected ranges: mean P-tau217 was 2.2 (SD 2.2) pg/mL, GFAP 1.3 (SD 1.2) pg/mL, and NfL 2.9 (SD 3.7) pg/mL. There were no appreciable sex differences in unadjusted P-tau217 or NfL levels; females had marginally higher GFAP on average.

**Table 1.** Summaries of analytic sample.

|  | Male<br>N=2500 | Female<br>N=2973 | Total<br>N=5473 |
| --- | --- | --- | --- |
| **Childhood poverty** |  |  |  |
| No | 2298 (91.9%) | 2753 (92.6%) | 5051 (92.3%) |
| Yes | 202 (8.1%) | 220 (7.4%) | 422 (7.7%) |
| **P-tau217** | 2.2 (2.1) | 2.1 (2.2) | 2.2 (2.2) |
| **NfL** | 3.0 (3.8) | 2.8 (3.5) | 2.9 (3.7) |
| **GFAP** | 1.2 (1.3) | 1.3 (1.0) | 1.3 (1.2) |
| **Age** | 66.5 (8.6) | 65.4 (8.7) | 65.9 (8.7) |
| **College education** |  |  |  |
| Less than coll. | 1556 (62.2%) | 1886 (63.4%) | 3442 (62.9%) |
| College | 944 (37.8%) | 1087 (36.6%) | 2031 (37.1%) |
| **Marital status** |  |  |  |
| Single | 167 (6.7%) | 142 (4.8%) | 309 (5.6%) |
| Married/Partnered | 1943 (77.7%) | 2066 (69.5%) | 4009 (73.3%) |
| Sep/div/widowed | 390 (15.6%) | 765 (25.7%) | 1155 (21.1%) |
| **Bottom wealth quartile** |  |  |  |
| No | 2478 (99.1%) | 2934 (98.7%) | 5412 (98.9%) |
| Yes | 22 (0.9%) | 39 (1.3%) | 61 (1.1%) |
| **Country** |  |  |  |
| Germany | 967 (38.7%) | 1079 (36.3%) | 2046 (37.4%) |
| Sweden | 708 (28.3%) | 893 (30.0%) | 1601 (29.3%) |
| Denmark | 825 (33.0%) | 1001 (33.7%) | 1826 (33.4%) |

In pooled Poisson regressions adjusting for demographics and socioeconomic factors, childhood poverty was significantly associated with NfL but not with GFAP or P-tau217 (Table 2). Compared to the non-poor, those who grew up poor had higher coefficient of NfL (*β* = 1.66, *p* < 0.001). The marginal/predicted values show that the magnitude of the NfL difference was marked: childhood-poor participants had on average 10.3 pg/mL (versus 2.0 pg/mL for the non-poor), reflecting a considerable increase relative to the sample mean (2.9 pg/mL). By contrast, no significant differences in P-tau217 or GFAP were observed.

**Table 2.** Coefficient estimates of degenerative biomarkers on childhood poverty, controlling for other factors (pooled sample)

|  | NfL |  |  | GFAP |  |  | P-tau217 |  |  |
| --- | --- | --- | --- | --- | --- | --- | --- | --- | --- |
|  | coef | s.e. | p | coef | s.e. | p | coef | s.e. | p |
| Constant | -1.37 | 0.43 | <0.001 | -1.58 | 0.24 | <0.001 | 0.92 | 0.24 | <0.001 |
| Childhood poor | 1.66 | 0.11 | <0.001 | -0.07 | 0.06 | 0.27 | -0.08 | 0.07 | 0.24 |
| Sex, Female | -0.15 | 0.10 | 0.14 | 0.06 | 0.06 | 0.34 | -0.06 | 0.06 | 0.32 |
| Age | 0.03 | 0.01 | <0.001 | 0.03 | 0.00 | <0.001 | 0.00 | 0.00 | 0.81 |
| College | -0.13 | 0.10 | 0.19 | 0.03 | 0.06 | 0.65 | -0.02 | 0.07 | 0.73 |
| Single | 0.02 | 0.18 | 0.93 | 0.05 | 0.12 | 0.67 | 0.02 | 0.09 | 0.87 |
| Partner/married | -0.06 | 0.16 | 0.70 | 0.01 | 0.07 | 0.89 | -0.10 | 0.07 | 0.15 |
| Bottom wealth quartile | -0.07 | 0.25 | 0.77 | -0.08 | 0.18 | 0.63 | -0.06 | 0.14 | 0.66 |

### Neurodegenerative biomarker age profiles by country

Figure 1 presents the adjusted marginal/predicted age profiles of each biomarker as a function of age (50–85+ years) in Germany, Sweden, and Denmark, comparing those with childhood poverty (solid lines) and without (dashed lines).

**Figure 1.**
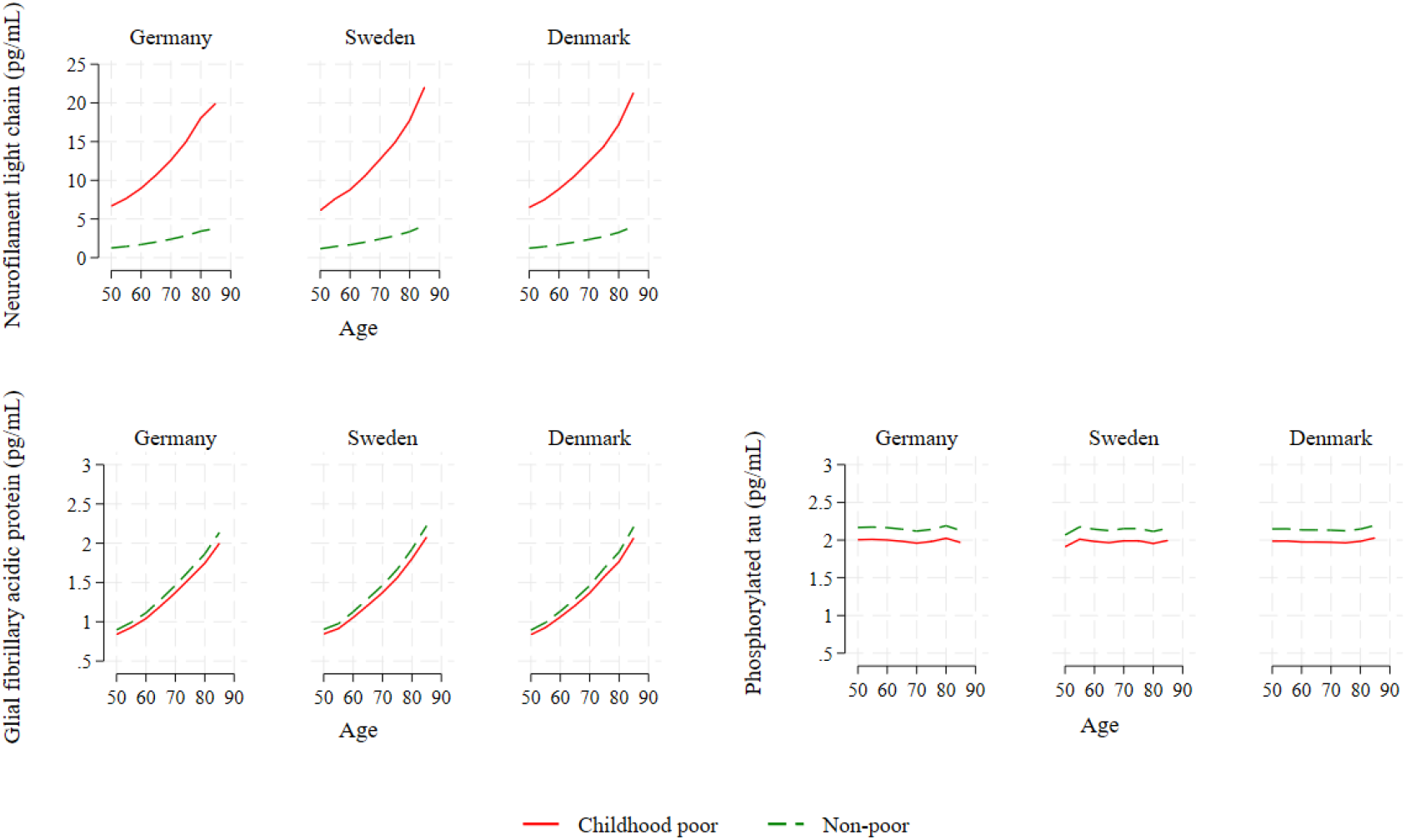
Marginal plots for each country, distinguishing the childhood poor (solid lines) from non-poor (dashed lines)–. neurofilament light chain (NfL, top left), glial fibrillary acidic protein (GFAP, bottom left), phosphorylated tau at threonine 217 (P-tau217, bottom right).

#### Neurofilament light chain (NfL)

The marginal predictions for NfL show a clear and consistent separation between those who were raised under poor material conditions and those non-poor. In all three countries, predicted NfL rises with age for both groups, but the increase is markedly steeper among the childhood-poor, producing a widening gap at older ages. In fact, the middle aged among the childhood poor had already had poorer start than the oldest old among the childhood non-poor. This pattern aligns with the pooled Poisson model in which childhood poverty is strongly associated with higher NfL net of age, sex, education, marital status, wealth in any country. The age-profile in the plots therefore suggests that the childhood-poverty association is not merely a level shift at a given age; rather, the disparity becomes more pronounced at older ages, consistent with a life-course imprint that is expressed as elevated neuro-axonal injury in later life.

#### Glial fibrillary acidic protein (GFAP)

In contrast, the marginal plots for GFAP show substantial overlap between childhood-poor and non-poor cohort members in each country. Predicted GFAP increases with age in each country, indicating a strong age gradient, but differences by childhood poverty status are small and not significant. The regression and marginal predictions indicate that childhood poverty is not associated with later life astrocytic activation/neuroinflammatory signalling as captured by DBS-based GFAP, whereas age is.

#### Phosphorylated tau (p-tau217)

The marginal plots for p-tau217 are largely flat across age and show non-significant separation by childhood poverty status in each country. This again is consistent with the pooled models, which do not detect an adjusted association between childhood poverty and p-tau217.

The combined evidence from Table 2 and the marginal predictions therefore suggests that the childhood-poverty signal in this study is specific to NfL rather than extending to GFAP and p-tau217 in a general population sample. Overall, the marginal plots reinforce the main message from the adjusted regressions: childhood poverty is associated with later life neurodegeneration as measured by NfL, while GFAP and p-tau217 show no corresponding pattern. Importantly, these contrasts are observed consistently across Germany, Sweden, and Denmark, supporting the cross-country robustness of the NfL association in the SHARE biomarker study.

## Discussion

Our finding that NfL concentrations, indicating neuroaxonal damage, are shaped by childhood poverty echoes the emerging literature on the life course shaping of health especially cognitive health.^2,6,7,12^ Now we know childhood poverty also shapes brain health. This association is observed consistently across the three European countries, suggesting that childhood poverty exerts a durable influence on brain ageing irrespective of specific histories and health systems. This finding stands in contrast to recent literature on the life course shaping of health which suggests that Sweden is exceptional even within Europe.^16,17^

A particularly notable finding is the magnitude of this association when expressed in age-equivalent terms. Individuals in midlife who experienced childhood poverty exhibit NfL concentrations that exceed those observed among the oldest old^18^ who were raised in non-poor circumstances. This contrast corresponds to a difference of approximately three decades of biological ageing. Such a pattern indicates that childhood poverty is associated not merely with incremental elevations in neurodegenerative biomarkers, but with a substantial shift in the trajectory of axons integrity across the life course. The implication is that exposure to poverty early in life may accelerate neurobiological ageing to an extent comparable to several decades of chronological ageing. We are told that the childhood poor in America age epigenetically faster,^2^ now we find that their similarly poor peer in Europe age neurologically faster too.

Equally noteworthy is the variation across the age profiles of the biomarkers: NfL has an fan shape, GFAP has an incline shape, P-tau217 has a flat shape. Recall that exactly the same weighted Poisson model was fitted to each. Yet the age profiles are very different. This difference is consistent with the different roles for each biomarker in brain function.

The magnitude and persistence of the association between childhood poverty and later life NfL concentrations are consistent with the hypothesis of biological embedding. Early life deprivation may induce long-term alterations in stress-response systems, inflammatory pathways, and metabolic regulation, which cumulatively contribute to axonal injury. Conditions of childhood poverty, including constrained access to adequate nutrition, greater exposure to environmental hazards, and limited opportunities for cognitive and social stimulation, may further compound these effects. The consistency of the association across national contexts suggests that these mechanisms operate through fundamental biological processes that are robust to variation in histories, welfare regimes and health systems.

These findings extend previous work linking childhood poverty to poorer cognitive function.^2,12^ By demonstrating an association with circulating NfL, the current study identifies a plausible biological substrate that may underlie these epidemiological relationships. NfL is recognised as a sensitive marker of neuroaxonal damage across a range of neurodegenerative conditions, and elevations in NfL have been shown to precede clinical manifestations of cognitive decline in this sample and beyond.^3,4^ The observed association therefore supports a life-course model in which childhood poverty contributes to subclinical neurodegenerative processes decades before the onset of overt disease.

Other biomarkers of neurodegeneration, including phosphorylated tau and glial fibrillary acidic protein, were also examined. While these markers reflect distinct pathological processes, the most consistent and pronounced association was observed for NfL. This may reflect the sensitivity of NfL to cumulative and non-specific neuroaxonal damage arising from exposures associated with childhood poverty. The pattern observed here is consistent with the interpretation that NfL captures the aggregate burden of life-course insults affecting brain integrity.

The methodological contribution of the study is nonetheless important. The harmonised analysis of multiple population-based cohorts, each with comparable biomarker measurements and retrospective life-history data, enhances the generalisability of the findings. Cross-national comparability reduces the likelihood that the observed associations are artefacts of specific sampling frames or institutional settings.

Several limitations should be considered. Residual confounding cannot be excluded despite extensive adjustment, particularly with respect to unobserved early life conditions. In addition, biomarker measurements were cross-sectional, precluding direct assessment of within-individual trajectories over time. Nevertheless, the strength and consistency of the association, including its magnitude relative to chronological ageing, support the interpretation of a substantive life-course effect.

The findings have important implications for both research and policy. The identification of childhood poverty as a determinant of neurodegenerative processes in later life highlights the potential for early life interventions to yield long-term benefits for brain health. Reductions in childhood poverty may therefore contribute not only to immediate improvements in well-being, but also to the prevention or delay of neurodegenerative disease at older ages. Future research should prioritise longitudinal designs incorporating repeated biomarker measurements to examine whether improvements in early life conditions are associated with slower trajectories of neurodegeneration.

In conclusion, exposure to childhood poverty is associated with markedly elevated NfL concentrations in later life across diverse European populations. The magnitude of this association, several decades of additional biological ageing, emphasises the importance of childhood poverty in shaping neurobiological ageing. The study also demonstrates the value of harmonised cross-national analyses integrating life histories and biomarker data, providing a foundation for future research on the life course shaping of brain health.

## Data Availability

SHARE is freely available to researchers after registration at www.share-project.org.

## Acknowledgements

We thank the cohort members for providing information, time and in 2015 blood biomarkers too, as well as the generous funding bodies over many decades. SHARE: This paper uses data from SHARE Waves 3, 7 and 9 (DOIs: 10.6103/SHARE.w3.800, 10.6103/SHARE.w7.800, 10.6103/SHARE.w9ca800) see Börsch-Supan et al.^10^ for methodological details. The SHARE data collection has been funded by the European Commission, DG RTD through FP5 (QLK6-CT-2001-00360), FP6 (SHARE-I3: RII-CT-2006-062193, COMPARE: CIT5-CT-2005-028857, SHARELIFE: CIT4-CT-2006-028812), FP7 (SHARE-PREP: GA N°211909, SHARE-LEAP: GA N°227822, SHARE M4: GA N°261982, DASISH: GA N°283646) and Horizon 2020 (SHARE-DEV3: GA N°676536, SHARE-COHESION: GA N°870628, SERISS: GA N°654221, SSHOC: GA N°823782, SHARE-COVID19: GA N°101015924) and by DG Employment, Social Affairs & Inclusion through VS 2015/0195, VS 2016/0135, VS 2018/0285, VS 2019/0332, and VS 2020/0313. Additional funding from the German Ministry of Education and Research, the Max Planck Society for the Advancement of Science, the U.S. National Institute on Aging (U01_AG09740-13S2, P01_AG005842, P01_AG08291, P30_AG12815, R21_AG025169, Y1-AG-4553-01, IAG_BSR06-11, OGHA_04-064, HHSN271201300071C, RAG052527A) and from various national funding sources is gratefully acknowledged (see www.share-project.org).

## Data availability

SHARE is freely available to researchers after registration at www.share-project.org.

